# Selective visuoconstructional impairment following mild COVID-19 with inflammatory and neuroimaging correlation findings

**DOI:** 10.1101/2022.03.16.22272467

**Authors:** Jonas Jardim de Paula, Rachel Elisa Rodrigues Pereira de Paiva, Nathália Gualberto Souza e Silva, Daniela Valadão Rosa, Fabio Luis de Souza Duran, Roney Santos Coimbra, Danielle de Souza Costa, Pedro Robles Dutenhefner, Henrique Soares Dutra Oliveira, Sarah Teixeira Camargos, Herika Martins Mendes Vasconcelos, Nara de Oliveira Carvalho, Juliana Batista da Silva, Marina Bicalho Silveira, Carlos Malamut, Derick Matheus Oliveira, Luiz Carlos Molinari, Danilo Bretas de Oliveira, José Nélio Januário, Luciana Costa Silva, Luiz Armando De Marco, Dulciene Maria de Magalhães Queiroz, Wagner Meira, Geraldo Busatto, Débora Marques Miranda, Marco Aurélio Romano-Silva

**Author notes:** **Corresponding author:** Prof. Marco A. Romano-Silva. Faculdade de Medicina da UFMG. Av Alfredo Balena 190. 30130-100. Belo Horizonte. Brazil. Contact number: 55-31-9-8399-7175.

## Abstract

People recovered from COVID-19 may still present complications including respiratory and neurological sequelae. In other viral infections, cognitive impairment occurs due to brain damage or dysfunction caused by vascular lesions and inflammatory processes. Persistent cognitive impairment compromises daily activities and psychosocial adaptation. Some level of neurological and psychiatric consequences were expected and described in severe cases of COVID-19. However, it is debatable whether neuropsychiatric complications are related to COVID-19 or to unfoldings from a severe infection. Nevertheless, the majority of cases recorded worldwide were mild to moderate self-limited illness in non-hospitalized people. Thus, it is important to understand what are the implications of mild COVID-19, which is the largest and understudied pool of COVID-19 cases. We aimed to investigate adults at least four months after recovering from mild COVID-19, which were assessed by neuropsychological, ocular and neurological tests, immune markers assay, and by structural MRI and ^18^FDG-PET neuroimaging to shed light on putative brain changes and clinical correlations. In approximately one-quarter of mild-COVID-19 individuals, we detected a specific visuoconstructive deficit, which was associated with changes in molecular and structural brain imaging, and correlated with upregulation of peripheral immune markers. Our findings provide evidence of neuroinflammatory burden causing cognitive deficit, in an already large and growing fraction of the world population. While living with a multitude of mild COVID-19 cases, action is required for a more comprehensive assessment and follow-up of the cognitive impairment, allowing to better understand symptom persistence and the necessity of rehabilitation of the affected individuals.

## INTRODUCTION

The severe acute respiratory syndrome coronavirus 2 (SARS-CoV-2) manifests itself as a mild respiratory tract infection in most individuals, leading to COVID-19 disease [1]. In some infected individuals, this can progress to deadly severe pneumonia and acute respiratory distress syndrome (ARDS). Nevertheless, the majority of cases recorded worldwide were mild to moderate self-limited illness in non-hospitalized people. In the beginning of the pandemic, the WHO–China Joint Mission on COVID-19 report 80% of the 55 924 patients with laboratory-confirmed COVID-19 in China to Feb 20, 2020, had mild-to-moderate disease, while 13.8% developed severe disease and 6.1% evolved to critical stage requiring intensive care [2, 3].

As of February 10th, 2022, there has been estimated more than 402 million confirmed cases of COVID-19, including more than 5.7 million deaths, reported to WHO. Among the notified cases, it is expected that more than 320 million (80%) have had mild to moderate COVID-19. Now, more than 24 months after the start of the events that overturned the health systems across the world, vaccines are being widely distributed (10,095,615,243 doses administered) but new worries are emerging.

Vaccine distribution worldwide is heterogeneous, so the emergence of new variants, more transmissible strains and less severe presentation has been the new status quo. These new variants have been more transmissible, leading to a sharp increase in cases in shorter periods and infecting a larger number of people with a less severe presentation [4]. However, due to the high number of infected people and high transmissibility, there is an increasing risk of reaching everyone and overwhelming the health system. Interestingly, the mild COVID-19 has been less studied than the moderate and severe forms. Thus, it is important to understand what are the implications of having mild COVID-19.

A recent concern is the long-lasting presentation of COVID-19 which has been identified in about 5% of COVID-19 infected individuals a month after infection, and in up to 2% after four months [5]. Symptoms include fatigue, headache, cognitive compromise, dyspnea, and anosmia [5]. The association with higher number of symptoms [6] and the slow decrease of people having lasting symptoms [5, 7] suggests that the pathophysiological drivers underlying the presence of symptoms might be transient, such as inflammatory response. However, recent data shows that mild COVID-19 is related to important long-lasting symptoms, including neurological and psychiatric manifestations, and persistent sequelae, in about 30% of those patients, with increasing prevalence in aged individuals [8]. In England, a modeling of health economic impact of the long-COVID symptoms, depending on how long we would stand in pandemic times, estimates the government willingness-to-pay cost could reach more than 32 billion pounds to avoid the potential 557,764 QALY’s loss in population [9].

Undoubtedly, beyond the COVID-19 pandemic obvious consequences, it also carries a significant psychological stressor with a tremendous impact on individuals, organizations, as well as social and economic groups. Death and insecurity about the future are powerful psychological stressors and social isolation results in loss of educational activities, structured work, and mental health problems [10]. Historical records suggest that previous influenza pandemics of the XVIII and XIX centuries were marked by increased incidence of neuropsychiatric syndromes, such as insomnia, anxiety, depression, mania, psychosis, suicide, delirium [11–13], and neuromuscular or demyelinating processes. These usually appear during the acute viral phase or at subsequent periods after infection, in recovered patients. As an example, lethargic encephalitis had a surge in cases following the Spanish flu of 1918 [14, 15]. In the XXI century, there were reports of neuropsychiatric sequelae, such as narcolepsy, convulsions, encephalitis, encephalopathy, Guillain-Barre syndrome (GBS) and other neuromuscular and demyelinating processes, associated with SARS-CoV-1, H1N1 and MERS-CoV, virus from the same genus of the actual SARS-CoV-2 [16–18]. Thus, there were plenty of reasons to assume long-lasting neuropsychiatric symptoms associated with COVID-19. Indeed, attention should be paid to the direct effects of coronavirus infection, and its accompanying intense immunological response on the central nervous system (SNC) in addition to the psychological stress associated with it.

Magnetic resonance imaging data in hospitalized COVID-19 patients especially in individuals with severe forms of COVID-19 [19] demonstrated brain lesions [20–22]. A multimodal study including neuropsychological evaluation, MRI and PET-CT imaging, 29 hospitalized patients were evaluated [23]. They observed a frontoparietal damage with a distinctive pattern of lesions from sepsis, without attentional and processing speed worsening and with persistent changes in language and visual testes up to a month of discharge [23]. [24] investigated individuals who recovered from suspected or confirmed COVID-19 and showed these patients had a worse performance on cognitive tests in multiple domains when matched with non-COVID-19 subjects, showing evident deficits even amongst those without severe disease [24]. In a preliminary study, we demonstrated important deficits in the visuospatial processing in around 25% of mild (not requiring hospitalization) COVID-19 patients [25]. Here, we report results of a prospective observational cohort study of individuals with mild COVID-19 cases. They were investigated using neuropsychological tests, PET-CT and MRI neuroimaging, and immune markers analysis aiming to shed light on the mechanisms of long standing symptoms and related findings.

## METHODS

### Research design and procedures

Initially, we assessed clinical status, mental health and history of neurological symptoms with online questionnaires and scales, using the REDCap platform. All included individuals had positive RT-PCR for COVID-19 and mild COVID-19 presentation. After answering the questions, participants were assigned to subsequent procedures in two different visits. On the first one, neuropsychological assessment, neurological examination and brain structural magnetic resonance imaging (MRI) was performed. On the second visit, blood was collected and 18-FDG-PET brain imaging was performed. Figure 1 shows the overall research design.

**Figure 1.**
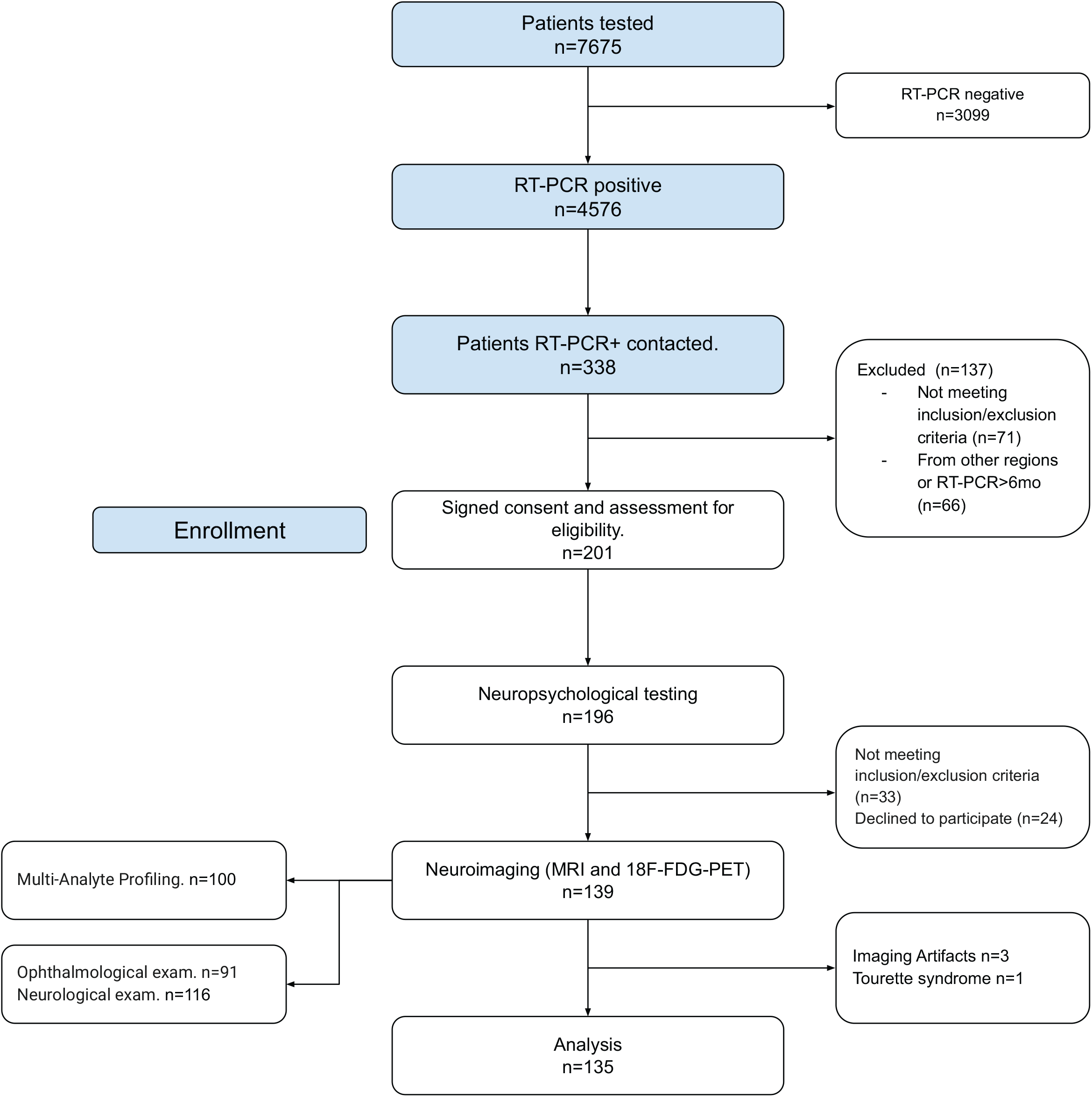
Research design and subsamples for each research procedure.

### Participants

The recruitment of participants was approved by the Institutional Review Board of Universidade Federal de Minas Gerais (UFMG) (CAAE 3768820.1.0000.5149) and followed international ethical standards. Written informed consent (digital and printed versions) was provided by all participants in the study. COVID-19 disease severity 1 and 2 according to the WHO clinical ordinal scale was referred to as mild COVID-19 [26]. Initial data collection was conducted using the Research Electronic Data Capture Platform (REDCap) and followed the Brazilian General Data Protection Law (LGPD) [27] for data collection, storage and use.

Volunteers were recruited through the NUPAD Center for Actions and Research in Diagnostic Support of Faculdade de Medicina at UFMG (FM-UFMG). We contacted a total of 338 patients with RT-PCR confirmed diagnosis of COVID-19. The average time between RT-PCR confirmation and inclusion was 4.35 (±2.45) months. Inclusion criteria consisted of adults with past mild COVID-19 cases in the last year, age between 18 and 60 years, and confirmed diagnosis of SARS-CoV-2 by RT-PCR. The following exclusion criteria were applied: self-reported history of autoimmune disease, previous chronic mental disorders or neurological diagnosis, history of recurrent infections, substance abuse, previous brain surgery, and endotracheal or orotracheal intubation during hospitalization/treatment of SARS-CoV-2. After the previous screening we assessed 196 COVID-19 patients. During our analysis we excluded 4 patients from the neuropsychology subsample. One had a non-disclosed diagnosis of Tourette Syndrome, two were psychologists with previous knowledge of the adopted cognitive tests and one individual was excluded due to the presence of incidental brain lesions detected at MRI scanning during the research. The final sample had 192 participants (Table 1). From this point on our sample was stratified in three subsamples (neuropsychology, neuroimage and immunology) in order to maximize the number of participants for each method adopted in the study.

**Table 1:** sociodemographic aspects of the COVID-19 patients (n=192)

| Sociodemographic measures |  |  | n | % |
| --- | --- | --- | --- | --- |
| Age (group) | 20 - 29 |  | 39 | 20% |
|  | 30 - 39 |  | 75 | 39% |
|  | 40 - 49 |  | 46 | 2% |
|  | 50 - 59 |  | 32 | 17% |
| Sex | Male |  | 55 | 29% |
|  | Female |  | 137 | 71% |
| Education (highest degree) | High |  | 65 | 34% |
|  | College |  | 127 | 66 % |
| SES (classification) | Low |  | 18 | 10% |
|  | Medium |  | 114 | 62% |
|  | High |  | 52 | 28% |
| Access to private healthcare | Yes |  | 133 | 72% |
| Hospitalization due to COVID-19 | Yes |  | 11 | 6% |
| Self-reported COVID-19 symptoms | Fever |  | 79 | 41% |
|  | Headache |  | 147 | 77% |
|  | Chills |  | 87 | 45% |
|  | Dry cough |  | 105 | 55% |
|  | Sore throat |  | 77 | 40% |
|  | Myalgia |  | 131 | 68% |
|  | Shortness of breath |  | 63 | 33% |
|  | Ageusia |  | 112 | 58% |
|  | Anosmia |  | 123 | 64% |
|  | Diarrhea |  | 72 | 37% |
|  | Nausea |  | 58 | 30% |
|  | Vomit |  | 30 | 16% |
|  | Other symptoms |  | 42 | 22% |

Neuropsychological tests were available for 191 participants (one patient was unable to perform the tests due to anxiety symptoms). Neuroimaging data was available for 166 participants - excluding five as previously mentioned - other 26 images were excluded due to technical problems during data acquisition (6 MRI datasets and 20 FDG-PET datasets), which led to a final subsample of neuroimage data of 135 participants. Lastly, immunological data was acquired for 100 participants which had both neuropsychological and neuroimaging data.

Participants’ sociodemographic and the self-reported symptoms during the COVID-19 infection characteristics are shown on Table 1. We found no statistically significant differences between the subsamples in terms of age (χ^2^=1.08, p=0.982), education (χ^2^=0.253, p=0.993), sex (χ^2^=0.06, p=0.970) or socioeconomic classification (χ^2^=1.09, p=0.895). A detailed description of each subsample is available at supplementary table 1.

### Psychiatric assessment

In the online REDcap formulary participants initially answered two questions regarding mental health: “Have you ever been diagnosed with a mental disorder” and if the answer is positive, “Does this disorder persist to this day?”. These two questions were the initial psychiatric screening used as exclusion criteria. Besides, even if the participant did not report previous or current mental disorder we adopted two screening measure to document possible psychiatric symptoms:

#### DSM-5 Self-Rated Level 1 Cross-Cutting Symptom Measure-Adult [28]

This is a self-reported measure of mental health based on the most recent classification of the American Psychiatric Association. The adult version consists of 23 questions of 13 psychiatric domains. We scored the scale according to the DSM-5 guidelines, and classified the participants in “normal” ou “in risk for mental disorder” in each of the 13 domains. We also created a general psychopathology measure using the total score of the scale, in which higher scores represent worse mental health.

#### Self Reporting Questionnaire - SRQ-20 [29]

The SRQ-20 is a brief scale for assessment of non-psychotic psychiatric symptoms. Its 20 questions refer mainly to symptoms of depression, anxiety and somatic disorders. Scores range from 0 to 20 and higher scores represent worse mental health. The cutoff 7 (“normal”) / 8 (“clinical”) shows a high sensitivity and specificity for mental disorders, and was used to classify our participants.

### Cognitive assessment

We worked with three different levels of cognitive assessment. In each we sought to investigate the frequency of cognitive deficits shown by our patients. For this purpose we adopted standardized psychometric measures previously validated for the Brazilian population, with normative data available in technical manuals, papers or book chapters.

To assess the subjective perception of cognitive change (worsening) we adopted the AD8 scale [30]. To assess cognitive deficits we used a brief but comprehensive neuropsychological test battery designed to assess memory, executive functions, language, visuospatial processing, psychomotor speed and attention.

#### AD8 Screening Questionnaire [30]

This is a brief measure designed to detect cognitive changes possibly related to neurocognitive disorders. The AD8 consists of 8 questions referring to different aspects of cognitive impairment, including problems of judgment, memory and impairment in activities of daily living. Although usually adopted in cases of Alzheimer’s disease, we adopted the test for self-report and its original instructions, asking the participant to report cognitive changes after the recovery of COVID-19. The cutoff score 1 (“normal”) / 2 (“impairment”) was adopted for the classification of our participants. The summed scored was also uses

#### The Rey-Osterrieth Complex Figure Test - ROCF [31]

This is a task in which examinees are asked to copy and recall a complex geometric figure. This task involves multiple cognitive processes including visuoconstruction, planning, visuospatial processing and memory. We used a copy, immediate recall (3 minutes) and delayed recall (30 minutes). Results from copy and each memory recall range from 0 to 36, higher scores represent better performance.

#### Modified version of the switching verbal fluency test - msVFT [32]

A variation of the traditional category fluency test designed to emphasize the cognitive flexibility assessment. We used the categories animals and then fruits, each in 60 seconds. Each correct word is scored as a point. The patient then is required to produce pairs of words (one animal and one fruit), focusing on the assessment of cognitive flexibility. For the latter procedure we score the total of correct pairs. Higher scores in this test represents better performance.

#### Trail Making Test [33]

A classical test designed to assess psychomotor speed, attention and executive functions. uThe version used is composed of two parts: “A” and “B”, in which the participant must visually search and connect numbers (A) and numbers-letters (B). The test is scored based on the total time dispensed in each part.

#### Five Point Test - (FPT) [34]

A test designed to evaluate non-verbal fluency and requires the participant to generate a different drawing by connecting five points (as in the face of a dice). This task measures the ability to initiate and sustain the productivity and self-monitoring, which is a way to assess the executive functions. Performance is analyzed based on the number of unique designs the participant is able to make.

#### Digit Span [35]

This is a task assessing operational memory and attention. First the participant repeats a crescent series of digits, starting from 2 up to 9 numbers. Following this initial step the participant is presented to the backward, similar to the first one but he must repeat digits in the inverse order.

#### The Logical Memory Test [36]

A memory test where a short story is read to the participant and he must remember as much information as possible, in an immediate trial (as soon as the reading is finished) and a delayed trial (about 30 minutes from the reading). Each trial is scored on 23 elements, and higher scores represent better performance.

### Neurologic and Ophthalmological exams

A detailed neurological exam encompassing mental status, cranial nerves, motor and sensory function, tendon reflexes, coordination, gait and stance was performed by a neurologist. A total of 116 patients underwent a thorough neurological evaluation by two neurologists (STC and HO). The routine ophthalmology examination was performed by a certified ophthalmologist to rule out possible ocular conditions that could interfere with the assessments: refraction (self refractor, static and dynamic pupils, visual acuity examination (Snellen’s chart), eye alignment and motility, pupil, visual confrontation field, examination of the external eye and eye attachments, previous segment - biomicroscopy with slit lamp, tonometry, examination of the posterior segment (biomicroscopy, indirect binocular ophthalmoscopy and indirect ophthalmoscopy).

### Neuroimaging

#### Magnetic resonance imaging (MRI)

Brain imaging acquisitions were performed for every patient on a 3.0 T MRI system (Skyra; Siemens, Erlangen, Germany) with a 20-channel receive head coil. The protocol included isotropic three-dimensional (3D) T1, T2 and T2-FLAIR sequences, at the Hermes Pardini Institute. T1-weighted spin-echo sequence, isotropic 3D T2-WI turbo spin-echo (SPACE), isotropic 3D fluid-attenuated inversion recovery (FLAIR), diffusion-weighted MRI (DW-MRI), and susceptibility-WI imaging (SWI). Gadolinium-contrast was not administered.

#### PET/CT

Resting-state 18F-FDG PET/CT brain images for patients were acquired in a GE Discovery 690 (GE Healthcare, Millwalke, WI, USA) PET/CT scanner. Blood glucose level was checked prior to FDG administration. Only patients with < 140 mg/dl were injected. Subjects had at least 6 hours of fasting before the exam. After an intravenous bolus injection of 0.09 mCi/kg of 18F-FDG, subjects rested for 50 min in a quiet and dark room with minimum stimuli. PET brain images were acquired subsequently, with an acquisition time of 10 min, and reconstructed using the OSEM (Ordered Subsets Expectation Maximization) algorithm. Attenuation correction was performed using the CT image.

#### LUMINEX immunoassay

Peripheral blood samples were obtained in EDTA vacutainer. Plasma samples were collected after centrifugation of venous blood (3000 g for 15 min at 4ºC), which was aliquoted and stored at -80ºC until the analysis. The analysis of plasma biomarkers including chemokines, inflammatory cytokines, regulatory cytokines and growth factors were carried out at Instituto René Rachou – Fiocruz using the MultiPlex kit Cytokine/Chemokine/Growth Factor Convenience 45-Plex Human ProcartaPlex™ (Thermo Fisher Scientific. USA. EPXR450-12171-901) according to the manufacturer’s instructions. The biomarker concentrations were determined according to standard curves using a 5-parameter logistic fit and the results were expressed as pg/mL.

### Data analysis

#### Neuropsychological data

For each neuropsychological test we used the available Brazilian normative data, stratified usually by age and/or education, to produce adjusted Z-scores. Measures scored in time units were inverted, so negative Z-scores indicated worse performance while positive indicated better performance. We then used typical criteria for cognitive difficulties (scores -1.5 standard-deviations below the population parameters) to classify the performance of each participant in each test as “normal” or “impaired”. This corresponds to nearly the 8th percentile. Cognitive deficits should have values significantly higher than this. To double-check our classification and reduce possible bias from normative data we compared a subset of our participants (n=49) were matched in terms of age, education and sex with a control group from another study, conducted before the COVID-19 pandemic, that was assessed using the same protocol. These controls were used in psychometric studies for these neuropsychological tests, previously conducted by our group and published in Malloy-Diniz and colleagues (2018) [37]. Each participant of this control sample had its neuropsychological tests scored according to the previous procedures. Chi-square and Independent samples t-tests were used to compare the neuropsychological data of COVID-19 patients and controls.

#### Processing and analysis of neuroimaging data

MRI brain scans were initially inspected by two experienced radiologists and 18F-FDG-PET scans by a nuclear physician and a radiologist. All valid FDG-PET and T1-MRI datasets were converted from DICOM format to NIfTI format using DCM2NII software (http://www.cabiatl.com/mricro/mricron/dcm2nii.html), and images were oriented manually to place the anterior commissure at the origin of the three-dimensional Montreal Neurological Institute (MNI) coordinate system. FDG-PET images were co-registered to the T1-MRI datasets of the same individual, using PMOD™ version 3.4 (PMOD Technologies Ltd., Zurich, Switzerland), and corrected for partial volume effects (PVE) to avoid the confounding influence of regional brain atrophy (Thomas et al, 2011) and high white matter uptake (Matsubara et al, 2016). PVE correction on the co-registered FDG-PET images was conducted through the Meltzer method [38], an optimized voxel-based algorithm that is fully implemented in PVElab software (http://pveout.ibb.cnr.it/PVEOut_Software.htm) [39].

Using Statistical Parametric Mapping (SPM), version 12 (Wellcome Trust Center of Neuroimaging, London, United Kingdom), running in MATLAB R2012a (MathWorks, Sherborn, Massachusetts), the T1-MRI datasets of each individual were segmented into gray matter, white matter and cerebrospinal fluid using the unified segmentation model [40]. Subsequently, a custom template was created using Diffeomorphic Anatomical Through Exponentiated Lie Algebra [41]. This template was normalized into MNI space, and these parameters were applied to the separated gray matter, white matter and PVE-corrected FDG-PET images in order to achieve spatial normalization to MNI space [40]. Finally, the spatially normalized gray matter, white matter and PVE-corrected FDG-PET images were smoothed with a Gaussian filter of 8-mm at FWHM.

Using the voxel-based morphometry (VBM) statistical approach, we investigated the presence of linear correlations between regional GM/WM volumes and scores on each neuropsychological test for which COVID-19 patients presented significantly poorer performance relative to controls. The general linear model was used, based on random Gaussian field theory [42], entering age, sex and education (years) as confounding variables. The total intracranial volume (TIV) was also included as an additional covariate, calculated using the MATLAB get_totals script (http://www.cs.ucl.ac.uk/staff/g.ridgway/vbm/get_totals.m). Only voxels with values above an absolute GM and WM threshold of 0.05 entered these VBM analyses.

The same voxelwise approach was used to investigate the presence of significant linear correlations between regional glucose metabolism (using the PVE-corrected FDG-PET datasets) and scores on any neuropsychological test(s) for which COVID-19 patients presented poorer performance as compared to controls. To ensure the FDG-PET analysis only included voxels mapping cerebral tissue, a default threshold of 0.5 of the mean tracer uptake inside the brain was selected. Global uptake differences between brain scans were adjusted using the “proportional scaling” SPM12 option, with age, sex and education included as confounding factors.

In all analyses above, resulting statistics at each voxel were transformed to Z-scores and displayed as SPMs into standardized space, at a threshold of p < 0.0005 (two-tailed), corresponding to a Z score > 3.29, and an extent threshold of 10 contiguous voxels. These maps were then inspected for the presence of significant voxel clusters of positive or negative correlations with neuropsychological test results.

#### LUMINEX data

The Shapiro-Wilk Test was performed to define the normality distribution of data, which indicated non-parametric distribution. Mann-Whitney U Test was employed and correlation analyses were performed through Spearman rank for non-parametric observations. Z-scores at the Rey-Osterrieth Complex Figure (ROCF) test were compared with the Kolmogorov-Smirnov statistical test. *P* < 0.05 was considered statistically significant. Tests were performed using SPSS 25.0 and GraphPad Prism.

Grouping was performed with the hierarchical clustering algorithm parameterized with Spearman’s correlation and complete linkage, implemented in GenePattern [43]. Biomarkers were also clustered with the same parameterization, based on their expression values in the individuals.

Functional enrichment analysis was performed with the Ingenuity Pathway Analysis (IPA) software (Qiagen). The interaction network was built from the list of differentially expressed biomarkers and using the Connect function restricted to interactions experimentally observed or predicted with high confidence. All other parameters were set to default. Next, the most connected and informative componentes of the Ingenuity Toxicity List (clinical pathology endpoints) and Canonical Pathways were added with the Overlay function.

## RESULTS

### Sociodemographic data and report of COVID-19 symptoms

Our sample (n=192) was predominantly female (n=71%), relatively young (on average 38.17±9.82 years), highly educated (66% with a college degree or post graduation) and average socioeconomic status according to Brazilian standards (62%). Regarding the COVID-19 infection 6% were referred to hospitalization during the acute phase of the disease. The most reported symptoms were headache (77%), myalgia (68%) and anosmia (64%).

### Mental health, ophthalmologic and neurological symptoms

About 8% of our sample has a history of mental disorders (n=15), mostly depression and anxiety disorders. According to the SRQ-20 screening 91 participants (48%) showed non-psychotic psychiatric symptoms. Similar results were seen in the self-reported DSM-5 screening where a relatively similar number of participants showed signs of depression (49%), anxiety (53%), anger (47%) and sleep disorders (50%). Other symptoms are shown in Supplementary Table 2. Although these values are considerably high they do not refer to the mental disorders per se, but a higher number of symptoms when compared to the general population. A total of 116 patients underwent a thorough neurological evaluation by two neurologists (SC and HO). Isolated and nonspecific findings were encountered in a few patients, such as optokinetic nystagmus, absence of ankle reflexes, indifferent plantar responses, decreased pinprick sensitivity on distal extremity of toes, global tendon hyperreflexia, unsustained ankle clonus, postural tremor and intention tremor.

**Table 2:** Neuropsychological impairment in COVID-19 patients (n=191)

| Test | Impairment<sup>1</sup> |  | Patients<sup>2</sup> | Controls<sup>2</sup> | t-test<sup>3</sup> |  |
| --- | --- | --- | --- | --- | --- | --- |
|  | n | % | M(SD) | M(SD) | p | d |
| Verbal Fluency (animals) | 16 | 8% | 19.35 (5.59) | 19.76 (4.79) | 0.699 | -0.08 |
| Verbal Fluency (fruits) | 5 | 3% | 17.00 (4.14) | 16.24 (3.88) | 0.354 | 0.19 |
| Switching fluency (pairs) | 5 | 3% | 17.00 (4.14) | 8.84 (1.68) | 0.820 | -0.05 |
| **ROCFT - Copy** | **48** | **24%** | **34.14 (2.95)** | **29.22 (4.41)** | **0.001** | **1.31** |
| **ROCFT - Immediate Recall<sup>4</sup>** | **9** | **5%** | **20.61 (6.19)** | **17.22 (5.42)** | **0.003** | **0.58** |
| **ROCFT - Delayed Recall<sup>4</sup>** | **14** | **7%** | **21.04 (5.98)** | **16.41 (5.28)** | **0.001** | **0.82** |
| Logical Memory - Immediate Recall | 17 | 9% | 10.69 (3.55) | 11.76 (2.26) | 0.184 | -0.27 |
| Logical Memory - Delayed Recall | 10 | 5% | 9.82 (3.92) | 10.27 (4.10) | 0.581 | -0.11 |
| Digit Span Forward | 0 | 0% | 51.16 (22.88) | 50.22 (26.42) | 0.851 | 0.04 |
| Digit Span Backward | 0 | 0% | 24.82 (16.45) | 28.51 (15.09) | 0.250 | -0.23 |
| Five Point Test (unique) | 0 | 0% | 29.08 (11.6) | 27.45 (12.47) | 0.505 | -0.23 |
| Trail Making Test A | 21 | 11% | 37.45 (14.08) | 39.41 (14.46) | 0.498 | -0.23 |
| Trail Making Test B | 19 | 10% | 96.37 (54.78) | 85.67 (43.38) | 0.263 | -0.23 |

1-Compared to Brazilian normative data; 2-Matched by age, education and sex (n=49 for each group); 3-Independent samples t-tests and effect size calculated by the Cohens's d equation; 4-This differences were not significant after controlling for the copy impairment in an analysis of covariance (ANCOVA) model. M: mean, SD: standard-deviation, ROCFT: Rey-Osterrieth Complex Figure Test

### Subjective cognitive complaints

Regarding cognitive changes after recovering from the COVID-19 51% of our sample reported daily problems with thinking/or memory, 39% problems with judgment and 34% in remembering appointments. Figure 2 shows the full description of subjective cognitive changes measured by the AD8 scale.

**Figure 2.**
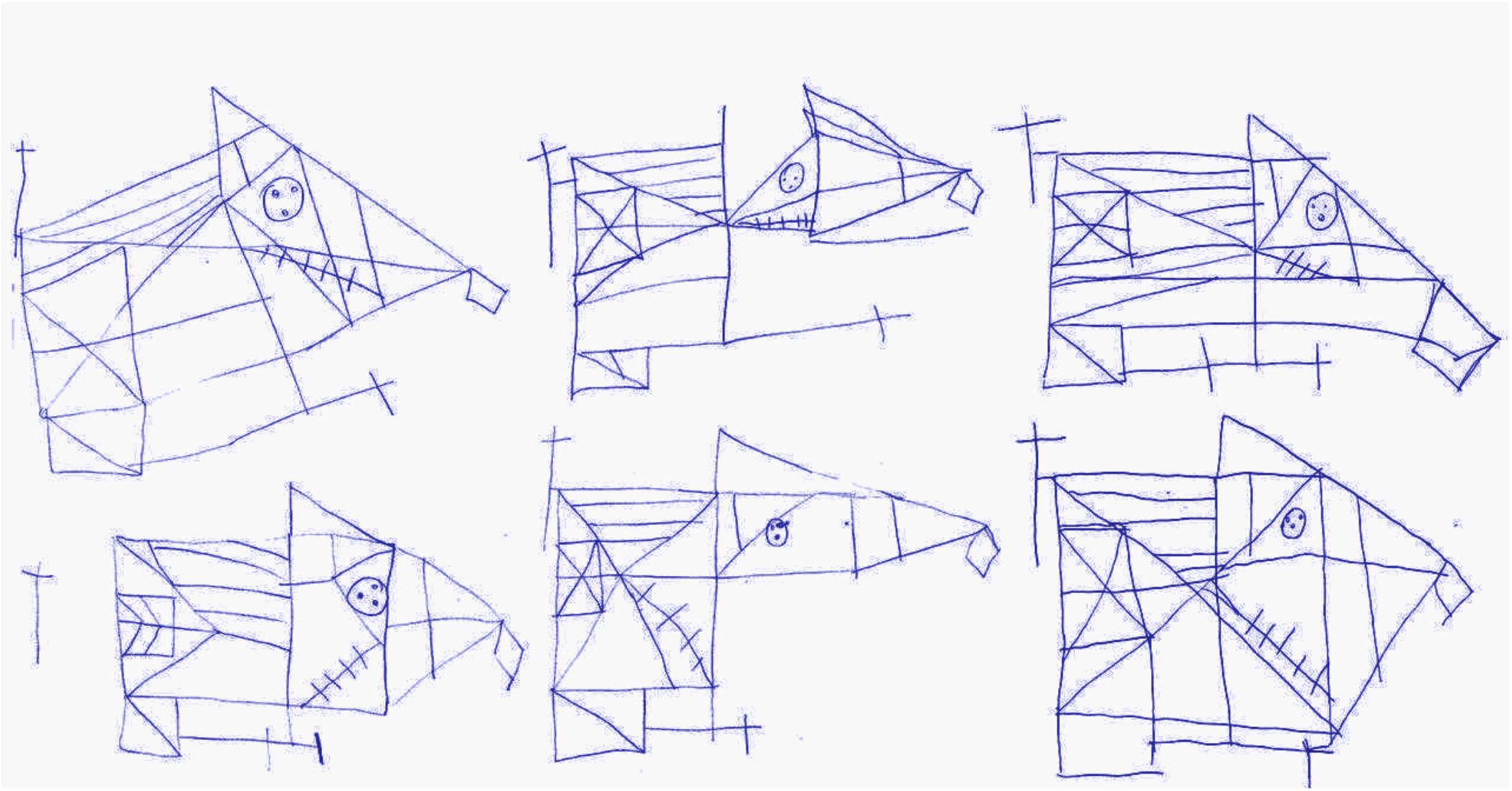
Examples of impaired performance in Rey-Osterrieth Complex Figure Test copy by COVID-19 patients.

### Neuropsychological assessment results

The frequency of cognitive impairment is shown on Table 2. We did not find significant differences in most of the neuropsychological tests, with frequencies of impairment around 8% (the expected for the criterion -1.5 standard-deviations below normative data). However, we found an atypically high rate of impairment in the copy ROCFT (26%). Matched controls showed about 6% of impairment in the same task.

When compared to the matched control sample we observed significant differences in this test. The copy trial was the most affected (t=6.40, p<0.001, d=1.31), while the immediate (t=2.88, p=0.003, d=0.58) and delayed recalls (t=4.06, p=0.001, d=0.82) showed less prominent differences. No other statistically significant difference was observed between patients and matched controls (p-values ranged from 0.184 to 0.870). Since previous studies suggest that the copy trial might significantly influence the memory recall (Shin et al., 2006), we then compared patients and controls in both memory trials using analysis of covariance (ANCOVA) controlling for the copy score, to investigate if the memory difficulties in the task were secondary to the visuospatial impairment. The analysis showed no differences in immediate (F=0.474, p=0.506) or delayed recall (F=0.219, p=0.650) after controlling for the copy score, which suggests a more specific impairment in the visuoconstructional processes of COVID-19 patients. Some examples of abnormal ROCFT copies are shown in Figure 2.

To further test if the cognitive deficits are restricted to the visuoconstructional processes we aim to test if these difficulties can be explained by symptoms of mental disorders or sociodemographic factors. We computed spearman rank-order correlations between neuropsychological tests Z-scores between these factors (Supplementary Figure 2). Most correlations were not statistically significant but we found a weak positive association of socioeconomic status and test performance (rho=0.290, p<0.01). However, this coeficiente was relatively similar in most neuropsychological tests, and did not seem a factor specifically related to the visuoconstructional measure. Even when patients and matched-controls were compared covariariating socioeconomic status the difference remained significant (p<0.001).

The neuropsychological profile of the COVID-19 showed a specific pattern of impairment in visuoconstructional processes, measured by the ROCF, in about 26% of the patients. To further investigate this deficit and its neurobiological correlates we stratified our sample in patients with and without impairment. Further analysis of immunological and neuroimaging data investigated the possible neurobiological factors associated with this deficit.

### Luminex multiplex assay findings

Neuroinflammation is characterized by activated microglia and reactive astrocytes in the brain parenchyma and increased levels of cytokines, chemokines, prostaglandins, complement cascade proteins, and reactive oxygen and nitrogen species (ROS/RNS), which can disrupt the blood brain barrier, allowing direct participation of the adaptive immune system.

Eleven biomarkers, namely LIF Interleukin 6 Family Cytokine (LIF), C-X-C motif chemokine ligand 10 (CXCL10), chemokine (C-C-motif) ligand 2 (CCL2), C-type lectin domain containing 11A (CLEC11A), C-C motif chemokine ligand 11 (CCL11), granulocyte-macrophage colony-stimulating factor (CSF), hepatocyte growth factor (HGF), interleukin 31 (IL31), interleukin 1 receptor antagonist (IL1RN), interleukin 10 (IL10) and nerve growth factor (NGF) were upregulated in the plasma of individuals with COVID-19 that showed visuospatial impairment in the copy of the Rey-Osterrieth Complex Figure Test when compared to patients without this deficit. Figure 3 shows the LUMINEX results.

**Figure 3.**
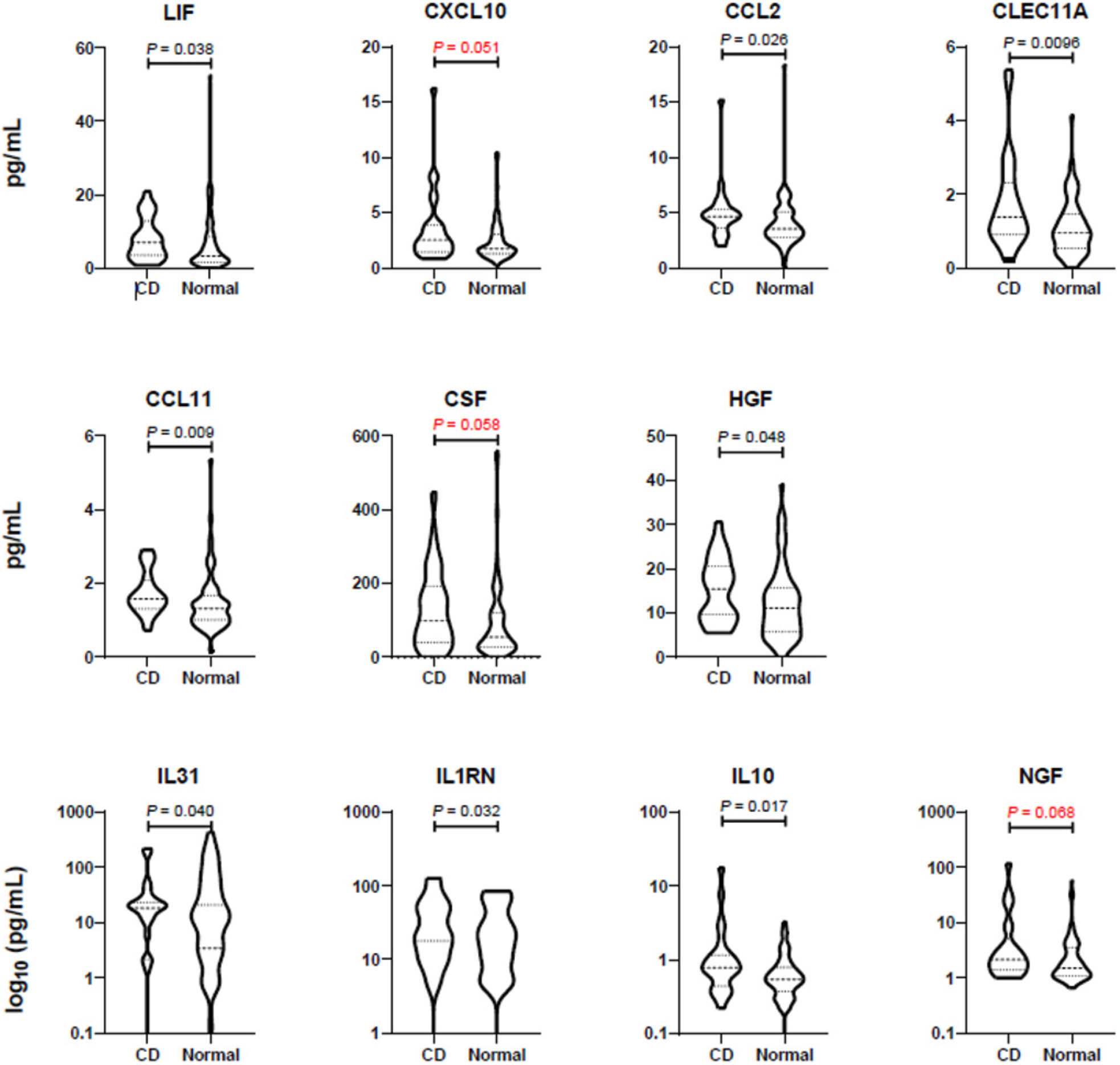
Differentially expressed plasma biomarkers associated with visuoconstructive deficit after mild COVID-19. Dashed lines = medians; dotted lines = quartiles. CD = visuoconstructive deficit. N for CD = 26; N for normal = 74.

Individuals were grouped based on the expression levels of 11 plasma biomarkers upregulated in those with visuoconstructive deficit when compared with those without deficit. In the hierarchical clustering, individuals were segregated in two main groups, one with low frequency of visuoconstructive deficit and relatively low levels of the 11 plasma biomarkers, the other with high incidence of deficit and higher levels of these proteins. This result suggests that lower expression of the 11 biomarkers is associated with protection against cognitive impairment (Figure 4A). Among the individuals with visuoconstructive deficit, at least four clusters were found (Figure 4B), indicating that the highly expressed 11 plasma biomarkers associated with cognitive impairment can be present at distinct combinatorial patterns. Individuals with visuoconstructive deficit from clusters 1 and 4 performed poorly at the ROCF test (Figure 4C).

**Figure 4.**
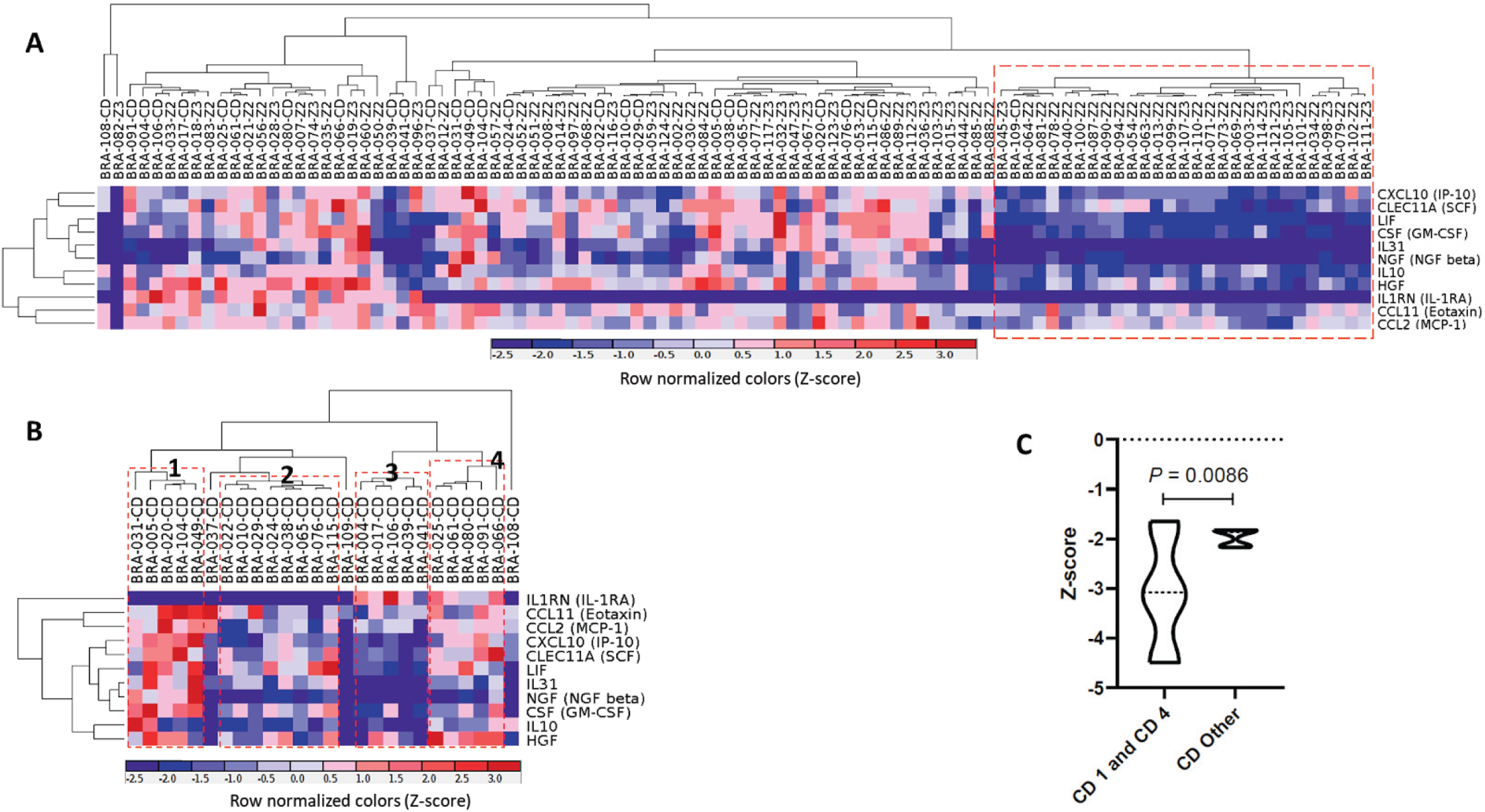
Hierarchical clustering of individuals with visuoconstructive deficit or normal outcome after mild COVID-19 and the differentially expressed plasma biomarkers. **A)** Hierarchical clustering of individuals with visuoconstructive deficit or normal outcome after mild COVID-19 and the differentially expressed plasma biomarkers. **B)** Hierarchical clustering of individuals with visuoconstructive deficit after mild COVID-19 and their upregulated plasma biomarkers. **C)** Comparison of Z-scores at the Rey–Osterrieth Complex Figure (ROCFT) test compared with Kolmogorov-Smirnov statistical test (interquartile).

Significant correlation was observed between the lowest values of the ROCF test and the highest plasma levels of SCF (c = 0.39, p = 0.048), CSF (c = 0.46, p = 0.020), HGF (c = 0.40, p = 0.041) and IL1RA (c = 0.59, p = 0.001), which was not observed in the group of individuals without visuoconstructive deficit (p ≥ 0,30 for all). The ROCF test values did not correlate with the age and the gender of the subjects as well as with the levels of the biomarkers (p ≥ 0,31 for all), but CCL11 increased plasma levels correlated with increasing age (c = 0.41, r = 0.001).

### Neuroimaging findings

We observed no structural changes in the MRI, and in particular no signs of thromboembolism, atrophy, acute encephalitis or leptomeningeal enhancement in any of the 135 patients. The VBM-based analysis of GM images showed no voxel clusters of significant positive or negative correlations with scores on the ROCF test, at the threshold of p<0.0005 and 10 contiguous voxels.

The analysis of WM images also showed no voxel clusters of significant positive correlation with scores on the ROCF test. Conversely, there was a large number of voxels (n=1,848) in which there were significant negative correlations between regional volumes and Z-scores on the ROCF test, indicating a widespread pattern of inverse relationship between test performance and WM volumes (see Figure 5). These voxels were aggregated in nine clusters (see Table 3), the largest of which (n=1,426 voxels) encompassing the subgenual portion of the corpus callosum and the cingulum on both hemispheres (Figure 5). The additional clusters involved WM portions of the inferior frontal gyrus and the fronto-occipital fasciculus bilaterally, as well as the right fusiform gyrus and the bilateral lingual gyri (Table 3).

**Table 3.** Significant correlations between performance on the Rey–Osterrieth Complex Figure Test and neuroimaging measurements of gray and white matter volumes (MRI) and glucose metabolism (FDG-PET).

| BRAIN REGION<sup>A</sup> | DIRECTION OF SIGNIFICANT CORRELATION | CLUSTER SIZE<sup>B</sup> | COORDINATES |  |  | PEAK Z-SCORE<sup>D</sup> |
| --- | --- | --- | --- | --- | --- | --- |
|  |  |  | x | y | z |  |
| GRAY MATTER VOLUME |  |  |  |  |  |  |
| No significant correlations. | - | - | - | - | - | - |
| WHITE MATTER VOLUME |  |  |  |  |  |  |
| Left and right genu of the corpus callosum, extending to the cingulum bundle | Negative | 1426 | -6 | 26 | -2 | 4.32 |
| Right fusiform gyrus | Negative | 93 | 32 | -22 | -26 | 4.01 |
| Right lingual gyrus | Negative | 127 | 30 | -44 | -8 | 3.84 |
| Right inferior frontal gyrus | Negative | 98 | 40 | 6 | 16 | 3.76 |
| Left lingual gyrus | Negative | 41 | -18 | -52 | 2 | 3.73 |
| Left inferior frontal gyrus | Negative | 20 | -34 | 30 | -2 | 3.48 |
| Left inferior fronto-occipital fasciculus | Negative | 15 | -24 | 4 | -8 | 3.47 |
| Left inferior fronto-occipital fasciculus | Negative | 15 | -32 | -10 | -8 | 3.44 |
| Right inferior fronto-occipital fasciculus | Negative | 13 | 32 | -10 | -8 | 3.43 |
| GLUCOSE METABOLISM (FDG-PET) |  |  |  |  |  |  |
| Left inferior temporal gyrus | Positive | 34 | -56 | -46 | -22 | 3.96 |
| Left inferior occipital gyrus (superior portion) | Positive | 53 | -48 | -68 | -16 | 3.92 |
| Right dorsal anterior cingulate gyrus | Negative | 69 | 8 | 16 | 42 | 4.61 |
| Right Rolandic operculum and opercular part of the inferior frontal gyrus | Negative | 57 | 52 | 8 | 12 | 4.48 |
| Right inferior occipital gyrus | Negative | 62 | 38 | -74 | -6 | 4.15 |
| Left calcarine and lingual gyri | Negative | 66 | -14 | -92 | -8 | 3.88 |
| Left superior frontal gyrus | Negative | 50 | -16 | 60 | -8 | 3.75 |
| Left inferior occipital gyrus (inferior portion) | Negative | 19 | -30 | -82 | -8 | 3.46 |
| Right medial frontal and orbital frontal gyri | Negative | 20 | 18 | 54 | -4 | 3.43 |

<sup>a</sup>For the analysis of white matter volumes, the brain regions where voxel clusters were located were identified according to the MRI Atlas of Human White Matter (Oishi et al., 2010). For the analysis of glucose metabolism, brain regions were identified according to the Automatic Anatomical Labeling Toolbox for SPM12 (Rolls et al., 2015); <sup>b</sup>Number of contiguous voxels in each cluster that surpassed the initial cutoff of $p \leq 0.0005$ ; <sup>c</sup>MNI coordinates of the voxel of maximal statistical significance within each cluster.

**Figure 5.**
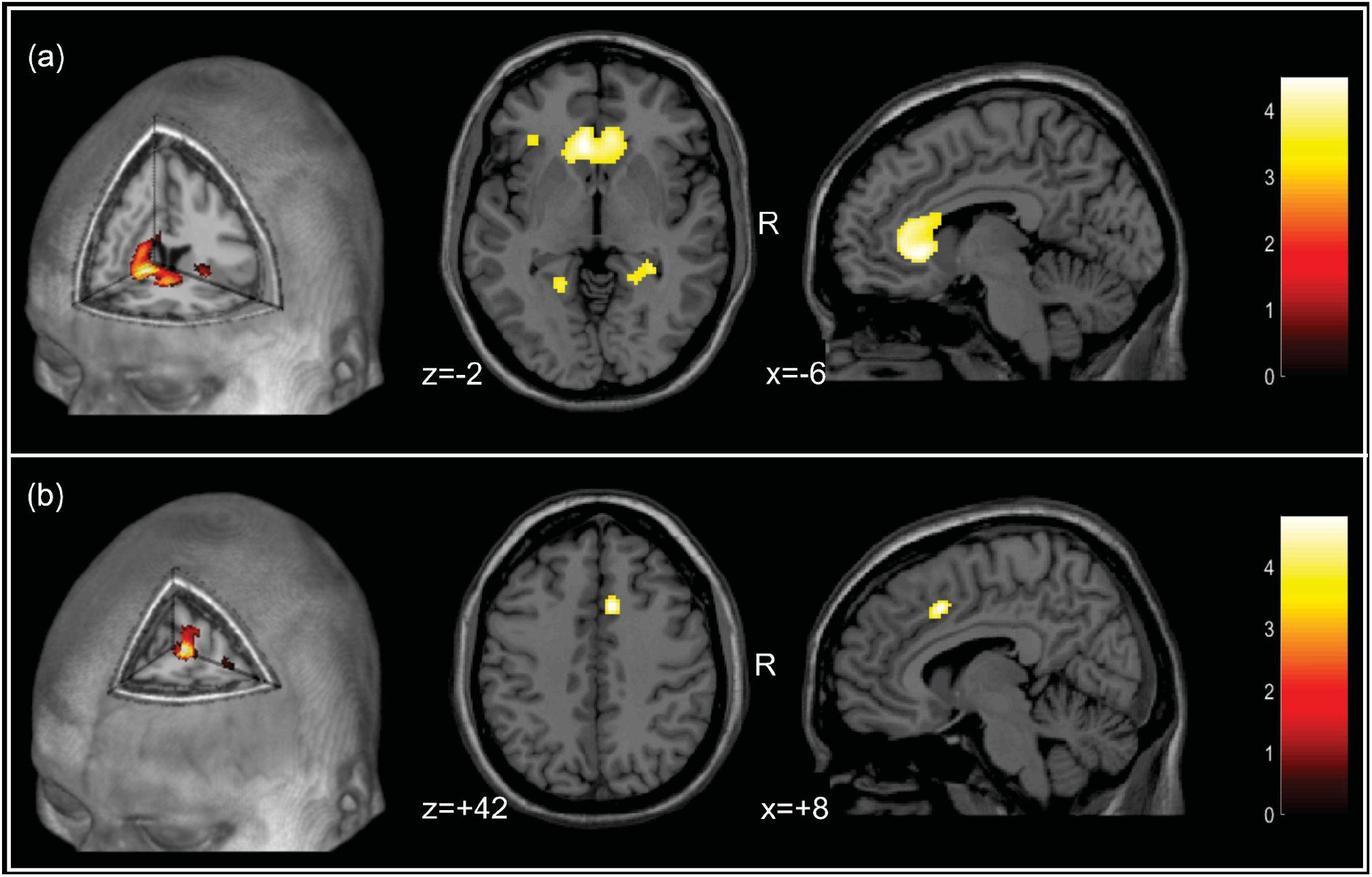
a) Findings of negative correlation between performance on the Rey-Osterrieth Complex Figure (ROCF) test and white matter volume (filtered at the Z>3.29 threshold). The foci show the peak of the greatest significance within the cluster (highlighted in yellow), located in the left and right genu of the corpus callosum, extending to the cingulum bundle; b) Findings of negative correlation between performance on the ROCF test and glucose metabolism (filtered at the Z>3.29 threshold). The foci show the peak of the greatest significance within the cluster (highlighted in yellow), located in the right dorsal anterior cingulate gyrus. The colored bar represents the T-value. Foci of significance were overlaid on axial brain slices spatially normalized into an approximation to the Talairach and Tournoux stereotactic atlas (Talairach and Tornoux, 1988). Abbreviations: R: right. Statistical details are provided in Table 3.

The analysis of FDG-PET images indicated nine clusters of voxels displaying significant correlations with Z-scores on the ROCF test (see Figure 5 and Table 3). Two of those were clusters of significant positive correlation, involving the left inferior temporal gyrus and the left inferior occipital gyrus (Table 3). The six clusters of significant negative correlation encompassed frontal (right dorsal anterior cingulate, Rolandic operculum and ventrolateral frontal cortices, and left superior lateral frontal cortex) and occipital regions (bilateral inferior occipital cortex, and left calcarine/lingual gyri (Table 3).

## DISCUSSION

In our sample of mild COVID-19 cases, we identified about half of participants with daily subjective cognitive problems and one-quarter of them with objective cognitive deficits, specifically a visuoconstructive impairment. Impaired performance on the ROCF copy, a classical measure of constructional praxis, was correlated with a pattern of increased brain white matter volume, with focal variations in brain glucose metabolism and altered expression of plasma immunomarkers.

Cognitive deficits following COVID-19 infection have been documented across studies of patients with different ages, disease severity and recovery time [44]. A smaller number of studies analyzed more specific cognitive functions, such as episodic memory, executive functions, verbal fluency [23] and sustained attention [45]. On the other hand, Mattioli and colleagues [46] reported no significant differences between patients and controls in neuropsychological tests, including the ROCF test (copy impairment was observed in only 5% of patients) four months after the infection. Altogether the studies suggest the presence of cognitive deficits after recovery from COVID-19. However, there are inconsistencies regarding affected cognitive functions and severity of deficits [47]. We observed significant cognitive impairment only in the ROCF, a drawing task test used to assess visuospatial abilities, executive functions and memory. The deficits observed in the ROCF could not be explained by socio-demographic factors, ophthalmologic deficits or psychiatric symptoms, suggesting cognitive deficit secondary to SARS-CoV-2 infection. Other factors which may influence performance, such as motor coordination, spatial neglect, visual attention, semantic knowledge, intelligence and executive functions were not likely to explain the observed difficulties, since we did not find any significant differences in other non-verbal (Trail Making Test and Five Points Test) and verbal tests (verbal fluency, digit span) also related to these processes. Drawing a complex figure involves different cognitive abilities related to perceiving, receiving, processing, storing and recalling visuospatial information, both regarding shape and position, and the planning and execution of the drawing per se. These processes involve multiple brain regions, including the occipito-parietal regions, the dorsal and ventral streams and connections with the cingulate, medial temporal and frontal cortices, integrating the perception and interpretation of the visual information with memory and executive systems [48–50]. Drawing tasks have been getting attention lately for their sensitivity to study visuospatial deficits, which were shown to be early biomarkers of neurodegenerative disorders, such as Alzheimer’s and Parkison’s disease [51–54].

The COVID-19 individuals investigated herein were often unable to produce a proper copy of Rey’s figure, and had difficulties in memory. Immediate and delayed recall seems to be secondary to the copy impairment. The lack of ability to assemble or organize parts into a whole object or figure is considered *constructional apraxia*, a neuropsychological syndrome which results in inability to accurately reproduce two-dimensional or three-dimensional visual models [55]. Constructional apraxia might follow acquired brain lesions or aging-related neurodegenerative diseases which affect the parietal or frontal lobes, but it is very uncommon in younger patients as the ones in our study, with a mean age of 38 years. Constructional apraxia is a sign of a divergence between the intended action and the actual performance, which may be seen in tests of free drawing or standardized tests of copy, including the ROCF. The performance on visuoconstruction and memory tests, such as the ROCF, are associated with different aspects of everyday life, including the capacity to learn, problem-solving skills, and activities of daily living [56]. There is a need for a more comprehensive assessment and follow-up of the visuoconstructive impairment, allowing to better understand symptom persistence and the necessity of rehabilitation of the affected individuals.

HCoVs have molecular structure and mode of replication similar to neuro-invasive animal coronaviruses [57] such as PHEV (porcine hemagglutination encephalitis virus) [58] and MHV strains (mouse hepatitis virus, MuCoV) [59], which can reach the CNS and induce different types of neuropathology. MHV is the best known coronavirus involved in short- and long-term neurological disorders [60, 61]. It is plausible to consider their involvement in neuropsychiatric symptoms and possible post-viral sequelae. SARS-CoV was detected in the serum and CSF of patients with persistent epilepsy and SARS [62]. The angiotensin-converting enzyme 2 (ACE2 or Ace2) has been identified as a primary entry receptor for SARS-CoV-2, as well as for SARS-CoV (Hoffmann et al.. 2020), indicating that SARS-CoV-2 may be able to infect the brain and result in CNS symptoms in patients with COVID-19 [63].

In other viral infections, such as in HTLV-1-infected individuals, a correlation between the proviral load in peripheral blood mononuclear cells was observed with brain white matter lesions, and deficits in tasks requiring integrity of subcortical activation, including constructive praxis [64]. As HTLV-1 seems not to be harbored by neurons *in vivo [65]*, persistent neuroinflammation was considered a possible explanation for white matter lesion and cognitive impairment in HTLV-1-infected patients [64]. The disruption of cytokines and chemokines signaling in the CNS has been postulated to contribute to the dysfunctional host-viral immune function and pathogenesis that occurs in inflammatory diseases such as HTLV-1 infection [66]. In contrast, neurons might become a target of SARS-CoV-2 infection, demonstrated in COVID-19 patient brain autopsies, mice brain in vivo and in vitro brain organoids [67]. In infections with influenza A virus (IAV), neuropsychiatric complications were reported after infection with neurotropic or non-neurotropic variants [68, 69], suggesting that viral infections can provoke neuroinflammation via peripherally-produced cytokines.

Microglia is the focal point of neuroinflammation, as it performs primary immune surveillance and activities similar to CNS macrophages. In infection or disease, the microglia cells become “activated” and function as inflammatory cell mediators. Depending on the context, the production of cytokines and chemokines can facilitate the recruitment of leukocytes to the brain [70]. Cytokines produced by the peripheral innate immune system can trigger a secondary neuroinflammatory response in the CNS, depicted by activation of microglia and production of proinflammatory cytokines (IL-1, IL-6, and TNF-alpha) [71–74]. Neuroinflammation was shown to severely affect cognition and behavior in animal models [69, 75–77]. It is mediated by the increase of cytokines and chemokines, reactive oxygen species (ROS) and second lipid messengers produced by astrocytes and microglia, endothelial and peripheral immune cells [78], with self-limitation after resolution [79, 80]. On the other hand, chronic neuroinflammation implies persistent activation of microglia and other immune cells in the CNS with potential damage [81–83]. Amplified or chronic inflammatory activation can lead to pathological changes and neurobehavioral complications, such as depression and cognitive deficits [77, 84, 85].

In this study, we measured peripheral blood immunity associated factors, which might partially correspond to the CNS findings discussed above. Among the 11 upregulated plasma biomarkers in individuals with visuoconstructive deficit, 10 composed a functional network based on experimental observation or high confidence computational predictions that they interact by up-or downregulating each other (Figure 6). These interactions are represented by the broken edges of the network connecting the nodes, which, in turn, represent the protein biomarkers. Furthermore, six of the 10 biomarkers in the network are related to the clinical pathology endpoints hepatic necrosis and five to cardiac necrosis, events that can result in the upregulation of these associated biomarkers. Finally, four biomarkers are components of the canonical “Neuroinflammation Signaling Pathway” and four of the “IL-17 Signaling pathway”.

**Figure 6.**
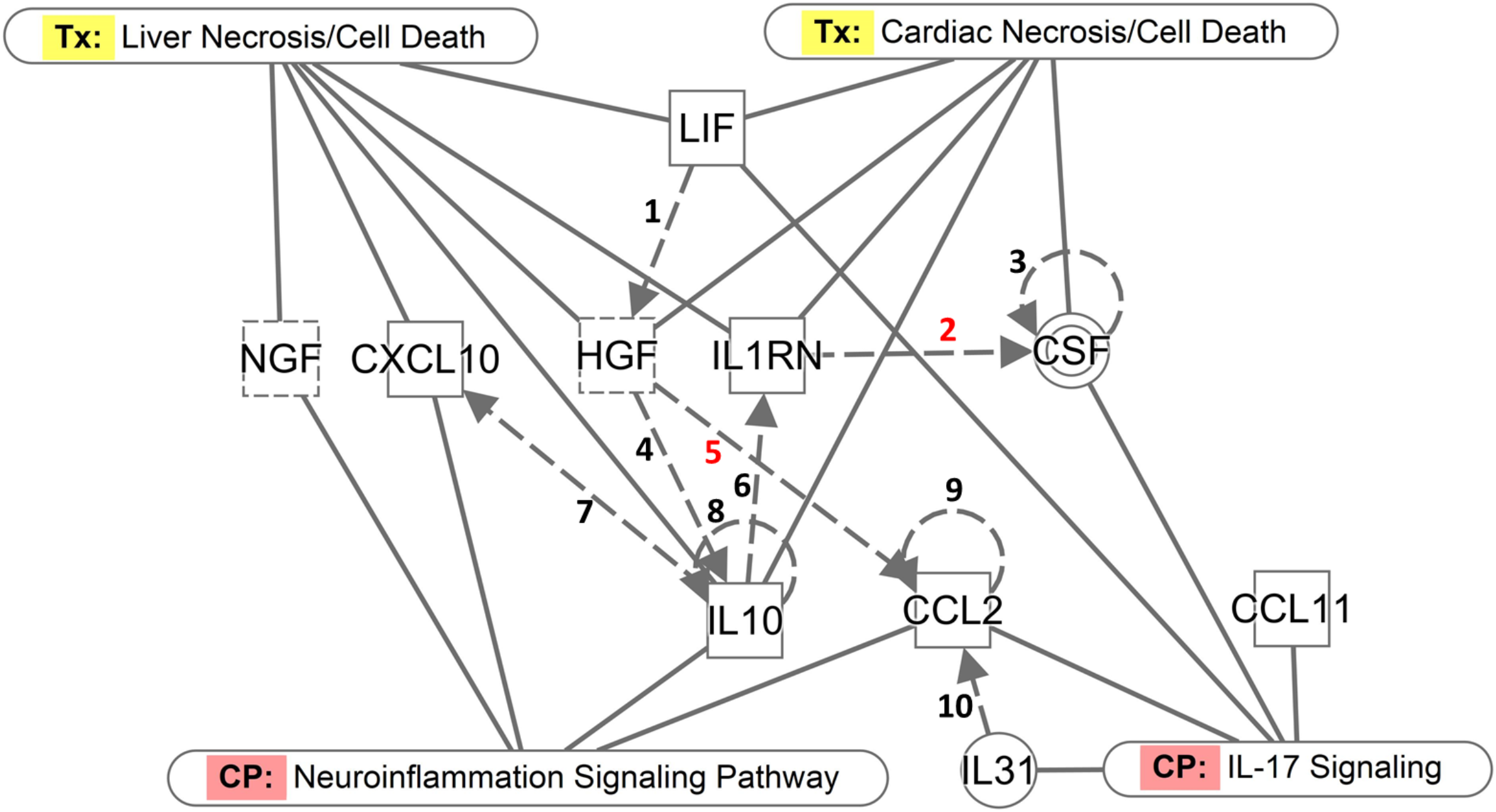
**-** Network of functional interactions between the differentially expressed plasma biomarkers (plain squares for cytokines or chemokines; dashed squares for growth factors, double circle for complex/group), canonical pathways (CP) and clinical pathology endpoints (Tx). (1) LIF protein increases expression of HGF mRNA [130]; (2) IL1RN protein decreases production of CSF protein [131]; (3) CSF protein increases its own dimerization [132]; (4) HGF increases secretion of IL10 protein [133]; (5) HGF protein decreases expression of CCL2 mRNA [134]; (6) IL10 protein increases expression of IL1RN mRNA [135]; (7) IL10 protein increases release of CXCL10 protein [136]; (8) IL10 protein increases expression of human IL10 mRNA [137]; (9) CCL2 mRNA is increased by CCL2 protein [138]; (10) IL31 protein increases expression of CCL2 mRNA [139, 140]).

The fact that 11 plasma biomarkers were associated with visuoconstructive deficit and 10 of them composed a functional network lend considerable support to the relevance of the immunological markers that may act either in the protection against or risk to cognitive impairment. Of note, the association was not only by a binary comparison, but also by comparing the degrees of the markers and of the visuoconstructive deficit.

Several upregulated biomarkers, identified in the present study, act on the central nervous system promoting protection or damage to the brain. HGF is a neuronal growth and survival factor capable of preventing neuronal death through its pro-angiogenic, anti-inflammatory and immunoregulatory activities and stimulating neuroregeneration by acting on neural stem cells. HGF also promotes synaptogenesis [86]. The presence of HGF might mean a sign of pre-conditions to recover after a multifaceted brain injury [86]. Another relevant biomarker found in the present study was NGF, which has anti-inflammatory properties in the central nervous system [87].

Other biomarkers, found to be related to ROCF-copy performance, act by promoting neuroinflammation. LIF is able to cross the blood-brain barrier and promote the induction of other pro-inflammatory cytokines in the central nervous system [88]. CXCL10 acts as an mediator for the activation and influx of leukocytes, such as T cells and others, in various inflammatory diseases of the central nervous system [89]. CSF is essential for the pathogenesis of experimental autoimmune encephalomyelitis (EAE), an animal model for multiple sclerosis, mediated by encephalitogenic T helper cells that produce IL-17 (Th17 cells) [90, 91]. Cytokine C-C motif chemokine 11 (CCL11, also known as eotaxin-1) can limit neurogenesis and contribute to cognitive impairment (Villeda et al., 2011). Serum levels of CCL11 has been an informative marker about prediction of worse and later functional outcome after ischemic stroke [92]. Elevated CCL11 levels were also found in the plasma of long-COVID patients with cognitive deficits compared to those without cognitive symptoms [93]. Notably, in accordance with the study of Villeda et al. (2011), an age-related increased CCL11 in the plasma of the patients with visuoconstructive deficit was observed in this study. Almost all biomarkers identified in the present study, with the exception of HGF and IL1RN, are components of the canonical pathways of IL-17 or Neuroinflammation signaling (CCL2 participates in both). In addition to the well-established association between neuroinflammation and cognitive impairment, the role of Th17 cells in brain diseases associated with this condition, such as multiple sclerosis, cerebral ischemia and Alzheimer’s disease, is also noteworthy. Th17 cells infiltrate the central nervous system where they induce direct brain cell damage or indirect effects mediated by disruption of the blood-brain barrier and neurovascular dysfunction [94]. Finally, strong evidence supports the association between CCL2 and cognitive impairment. In the nucleus of brain microglial cells, CCL2 decreases the activation of the protein GAD (aspartate 1-decarboxylase) [95]. GAD, in turn, decreases the density of GABAergic neurons [95] and increased GABAergic function in the prefrontal cortex impairs working memory [96]. Versace et al. reported that patients who had severe COVID-19 had reduced GABAergic inhibition in the primary motor cortex associated with fatigue and dysexecutive syndrome [97]. It is noteworthy that our cohort was made of individuals who had mild COVID-19. CSF assessment might be helpful to identify signs of neuroinfection, neuronal injury and degeneration. However due to the mild nature of symptoms of our participants, we did not collect CSF.

Plasma concentrations of almost all these biomarkers, with the exception of CCL2, CCL11 and IL31, are also upregulated in liver and heart injury, in which they play a protective and regenerative role. For example, IL1RN regulates cardiac remodeling by promoting the survival of cardiomyocytes in ischemic regions [98] and is also hepatoprotective [99]. IL-10 can have a dual role, compromising regeneration and increasing liver damage by blocking IL-6 production, or be hepatoprotective by inhibiting pro-inflammatory responses by TNF-alpha and lipopolysaccharide (LPS) [100], thus contributing to the healing of cardiac lesions [101]. We cannot rule out that individuals with visuoconstructive impairment post mild COVID-19 could also have some level of cardiac [102, 103] and/or hepatic damage [104]. Although our findings were from peripheral blood, there is a need to observe the long term correlates in liquor and other pathological specimens.

A recent study showed that mice mildly-infected with SARS-CoV-2 lost approximately one third of mature oligodendrocytes of the WM in the cingulum and corpus callosum, which was still present at 7-weeks post-infection [93]. WM-selective microglial reactivity was shown to inhibit neurogenesis, dysregulation of the oligodendrocytes and loss of myelin [105, 106] in both mice and humans following SARS-CoV-2 infection [93].

Our main MRI finding was an inverse relationship between ROCF test copy performance and WM volumes encompassing the subgenual portion of the corpus callosum and the cingulum on both hemispheres. The crossing over characteristics and the corpus callosum findings might have an important role in understanding the correlation with constructive apraxia [107]. Visuoconstructive deficit is a common finding in callosal ischaemic lesions [108] and agenesis [109]. Spatial abilities are expected to involve most prominently the right brain hemisphere, however some tasks have been shown to elicit bilateral brain activity [49]. Constructive apraxia has been associated with left- and right-hemisphere lesions, both in posterior and anterior regions, and their integration by white matter tracts.

Involvement of the right superior parietal lobe, angular gyrus, middle occipital gyrus have been previously associated with poorer performance in the ROCF copy task in pathological conditions, including vascular lesions [110], epilepsy [111], and traumatic brain lesions [112]. However, we found additional clusters involving WM portions of the inferior frontal gyrus and the fronto-occipital fasciculus bilaterally, as well as the right fusiform gyrus and the bilateral lingual gyri (Table 3).

Our main findings are an inverse association between ROCF test copy scores and the volume of brain white matter in different regions. Although seems paradoxical, since the correlation between brain volume and cognitive functioning is usually positive [113], according to a literature review of neuroimage findings in COVID-19 there is evidence that these patients might show an increased brain volume when compared to matched controls [114]. As reported by Lu and colleagues [115], recovered patients might show increased cerebral volume across different brain regions, including olfactory cortex, hippocampus, insula, left Rolandic operculum, left Heschl’s gyrus and right cingulate gyrus. Also, in some clinical conditions higher brain volume may not be beneficial for cognitive functioning, such as in patients with depression [116] and autism [117]. In diseases known to affect the white matter, such as multiple sclerosis, there are reports of transient increases in brain volume during periods of neuroinflammation relapse [118].

Brain metabolic changes have been documented across multiple studies of COVID-19 patients, although with inconsistent results [23, 119, 120]. Patterns of hypo-or hypermetabolism across different cortical and subcortical symptoms are related to neuropsychiatric symptoms of the disease, including anosmia [121], fatigue [122] and cognitive impairment [23]. Concerning resting brain glucose metabolism measured with ^18^FDG-PET, we found both positive and negative correlations with the ROCF-copy performance. Although the most significant cluster was of a negative correlation, located in the right dorsal anterior cingulate gyrus, we also found clusters of significant positive correlation, involving the left inferior temporal gyrus and the left inferior occipital gyrus. Changes in brain metabolism have been documented in COVID-19 patients, although its pattern is more inconsistent, and may be affected by the severity of disease, patients age and comorbidities [123]. Hosp and colleagues analyzed the pattern o covariance of PET-FDG brain images between COVID-19 patients and controls and reported several regions of cortical hypometabolism in the frontal and parietal lobes, as well as the caudate nuclei, and hypermetabolism in the white matter, cerebellum, brainstem and the mesial temporal lobe, and this pattern was predictive of cognitive deficits [23].

Our correlational approach highlighted regions in which brain metabolism (FDG-PET) or structural measures of white matter (MRI) were most strongly associated with ROCF copy performance. It should also be possible that regions functionally important to ROCF performance did not reach significance due to sample size and the lack of a control group.

Factors affecting enrollment into our prospective cohort study would not be expected to introduce selection bias, however, as our findings occurred when establishing the baseline, we must consider the potential to have a selection bias, causing an overestimate, since included individuals were invited for a neuropsychiatric study. Although the inclusion criteria was restrictive to previous mild cases of COVID-19 only, we must consider potential pulmonary or neurovascular damage, other mental disorders and psychosocial factors as important contributing factors related to the SARS-CoV-2 infection [124–127]. Nevertheless, considering the relatively high prevalence of cognitive deficits in neuroinfections [124, 125, 128] and their potential impact on daily living, it is paramount to keep investigating impairments caused by COVID-19.

For a while, there was not much attention directed to the large population affected by a mild COVID-19, since they apparently had recovered well. Even now, two years into the pandemic, it is still underinvestigated. It has been reported that home-isolated young adults (16-30 years old), with mild COVID-19, had persistent symptoms at 6 months, including fatigue, impaired concentration and memory problems [129]. Considering that we observed roughly 25% of our mild COVID-19 patients presenting visuoconstructive impairment, we can expect millions of people worldwide potentially suffering from this kind of deficit. It is imperative to approach populational samples to better understand the extension, causes and persistence of the dysfunction. Why is that so worrying? Constructive apraxia might stay undiagnosed without specific visuospatial testing, which does not mean it has no functional implications in daily life. Visuospatial ability is key to several daily living activities, such as driving, planning, drawing, to locate oneself in a place, and several occupations rely on good visuospatial perception, such as artists, surgeons, designers, pilots, among others. The functional adaptability must be evaluated to plan rehabilitation strategies. It is a deficit potentially reaching millions of mild diagnosed and undiagnosed COVID-19 cases.

A recent meta-analysis confirmed that fatigue, cognitive dysfunction and sleep disturbances are key features of post-COVID-19, but also highlighted that psychiatric manifestations such as sleep disturbances, anxiety, and depression increase significantly in prevalence over time [7]. Investigation of neuropsychiatric impacts and the pathophysiological drivers underlying the risk factors associated with COVID-19 are important in surveillance and in the development of evidence-based therapeutic strategies. Our findings provide evidence of putative neuroinflammatory burden, in an already large and growing fraction of the world population with mild COVID-19, which requires urgent confirmation, comprehension, and planning for mitigating actions.

## Data Availability

To protect the data privacy of the study participants, the dataset cannot be made publicly available. Specific data needed for reproducing results is available from the corresponding author upon reasonable request.

## CODE AVAILABILITY

Code needed for reproducing results is available from the corresponding author upon reasonable request.

### Competing Interests

The authors declare no competing interests.

Supplementary information is available at MP’s website.

## Acknowledgments

Coordenação de Aperfeiçoamento de Pessoal de Nível Superior - CAPES (processo 88881.504749/2020-01. 9951-Programa Estratégico Emergencial de Prevenção e Combate a Surtos. Endemias. Epidemias e Pandemias Edital de Seleção Emergencial I - Prevenção e Combate a Surtos. Endemias. Epidemias e Pandemias). Brazil; DMM, MARS, LADM, DMMQ, GB and WM are research fellows of the Conselho Nacional de Desenvolvimento Científico e Tecnológico (CNPq)-Brazil, FAPEMIG (APQ-00735-19), INOVA Fiocruz. The authors thank CDTN (agreement number 23072.047899/2018-13) for providing ^18^F-FDG doses; and Hospital Risoleta Tolentino Neves and Hospital Odilon Behrens.

