## Supplementary Information for "Selective visuoconstructional impairment following mild COVID-19 with inflammatory and neuroimaging correlation findings"

Running title: Cognitive impairment post-mild COVID-19

Jonas Jardim de Paula PhD[1,2], Rachel Elisa Rodrigues Pereira de Paiva[1], Nathália Gualberto Souza e Silva MSc[1], Daniela Valadão Rosa PhD[1], Fabio Luis de Souza Duran PhD[10], Roney Santos Coimbra PhD[12], Danielle de Souza Costa PhD[1], Pedro Robles Dutenhefner[1,8], Henrique Soares Dutra Oliveira[5], Sarah Teixeira Camargos MD, PhD[5], Herika Martins Mendes Vasconcelos MD[1], Nara de Oliveira Carvalho[7], Juliana Batista da Silva PhD[11], Marina Bicalho Silveira PhD[11], Derick Matheus Oliveira[8], Luiz Carlos Molinari MD[2], Danilo Bretas de Oliveira PhD[13], José Nélio Januário MD[5,7], Luciana Costa Silva Md, PhD[9], Luiz Armando De Marco MD, PhD[1,4], Dulciene Maria de Magalhães Queiroz MD, PhD[6], Wagner Meira Jr. PhD[10,14], Geraldo Busatto MD, PhD[10], Débora Marques Miranda MD, PhD[1,3,14] and Marco Aurélio Romano-Silva MD, PhD[1,2,14]

[1]Centro de Tecnologia em Medicina Molecular (CTMM), Universidade Federal de Minas Gerais (UFMG). Av Alfredo Balena 190. Belo Horizonte-MG, Brazil.

[2]Departamento de Saúde Mental; [3]Departamento de Pediatria; [4]Departamento de Cirurgia; [5]Departamento de Clínica Médica; [6]Departamento de Propedêutica Complementar; [7]Núcleo de Ações e Pesquisa em Apoio Diagnóstico (NUPAD). Faculdade de Medicina da Universidade Federal de Minas Gerais (UFMG).

[8]Departamento de Computação Científica, ICEX, Universidade Federal de Minas Gerais (UFMG), Belo Horizonte-MG, Brazil.

[9]Instituto Hermes Pardini, Rua Aimorés 66, Belo Horizonte-MG, Brazil.

[10]Departamento de Psiquiatria. Faculdade de Medicina da USP, São Paulo-SP, Brazil.

[11]UPPR, Centro de Desenvolvimento da Tecnologia Nuclear (CDTN), Belo Horizonte-MG, Brazil.

[12]Neurogenômica / Imunopatologia. Instituto René Rachou, Fiocruz, Belo Horizonte-MG, Brazil

[13]Faculdade de Medicina, Universidade Federal dos Vales do Jequitinhonha e Mucuri. Diamantina-MG, Brazil.

[14]Centro de Inovação em Inteligência Artificial para a Saúde (CIIAS-Saúde), Universidade Federal de Minas Gerais (UFMG), Belo Horizonte-MG, Brazil.

**Corresponding author:**

Prof. Marco A. Romano-Silva. Faculdade de Medicina da UFMG. Av Alfredo Balena 190. 30130-100. Belo Horizonte. Brazil. Contact number: 55-31-9-8399-7175.

### Supplementary Information

**Supplementary Table 1**. Full description of the study sample (n=191) and the subsamples which went to neuroimaging (n=135) and immunological (n=100) assessment.

| | | Neuropsychology | | Neuroimaging | | Immunological | |
|---|---|---|---|---|---|---|---|
| | | n | % | n | % | n | % |
| Age (group) | 20 - 29 | 39 | 20% | 25 | 19% | 23 | 23% |
| | 30 - 39 | 75 | 39% | 53 | 39% | 36 | 36% |
| | 40 - 49 | 46 | 24% | 34 | 25% | 23 | 23% |
| | 50 - 59 | 32 | 17% | 23 | 17% | 19 | 19% |
| Sex | Male | 39 | 26% | 32 | 25% | 25 | 25% |
| | Female | 111 | 74% | 96 | 75% | 76 | 75% |
| Education (highest degree) | High | 65 | 34% | 47 | 35% | 35 | 35% |
| | College | 127 | 66% | 88 | 65% | 66 | 65% |
| SES (classification) | Low | 18 | 10% | 18 | 13% | 12 | 12% |
| | Medium | 114 | 62% | 78 | 58% | 61 | 60% |
| | High | 52 | 28% | 38 | 28% | 28 | 28% |
| Private healthcare | Yes | 133 | 72% | 101 | 75% | 79 | 78% |
| Hospitalization | Yes | 11 | 6% | 9 | 7% | 5 | 5% |
| Self-reported COVID-19 symptoms | Fever | 79 | 41% | 59 | 44% | 42 | 42% |
| | Headache | 147 | 77% | 109 | 81% | 78 | 77% |
| | Chills | 87 | 45% | 68 | 50% | 53 | 52% |
| | Dry cough | 105 | 55% | 78 | 58% | 55 | 54% |
| | Sore throat | 77 | 40% | 55 | 41% | 44 | 44% |
| | Myalgia | 131 | 68% | 99 | 73% | 72 | 71% |
| | Shortness of breath | 63 | 33% | 46 | 34% | 33 | 33% |
| | Ageusia | 112 | 58% | 82 | 61% | 60 | 59% |
| | Anosmia | 123 | 64% | 89 | 66% | 68 | 67% |
| | Diarrhea | 72 | 38% | 56 | 41% | 40 | 40% |
| | Nausea | 58 | 30% | 45 | 33% | 35 | 35% |
| | Vomit | 30 | 16% | 27 | 20% | 22 | 22% |
| | Other | 42 | 22% | 38 | 28% | 32 | 32% |

**Supplementary Table 2**. Screening of psychiatric symptoms by the DSM-5 Self-Rated Level 1 Cross-Cutting Symptom Measure-Adult in a sample of 192 mild COVID-19 patients.

| **Symptom** | **n** | **%** |
|---|---|---|
| Depression | 94 | 49% |
| Anger | 90 | 47% |
| Mania | 51 | 27% |
| Anxiety | 101 | 53% |
| Somatic problems | 84 | 44% |
| Suicidal tougths | 25 | 13% |
| Psychosis | 20 | 10% |
| Sleep problems | 96 | 50% |
| Memory problems | 63 | 33% |
| Repetitive thoughts | 36 | 19% |
| Dissociation | 60 | 31% |
| Personality disorders | 46 | 24% |
| Substance use | 89 | 47% |

**Supplementary Figure 1. Heatmap correlation of psychiatric symptoms, sociodemographic factors and neuropsychological tests.**

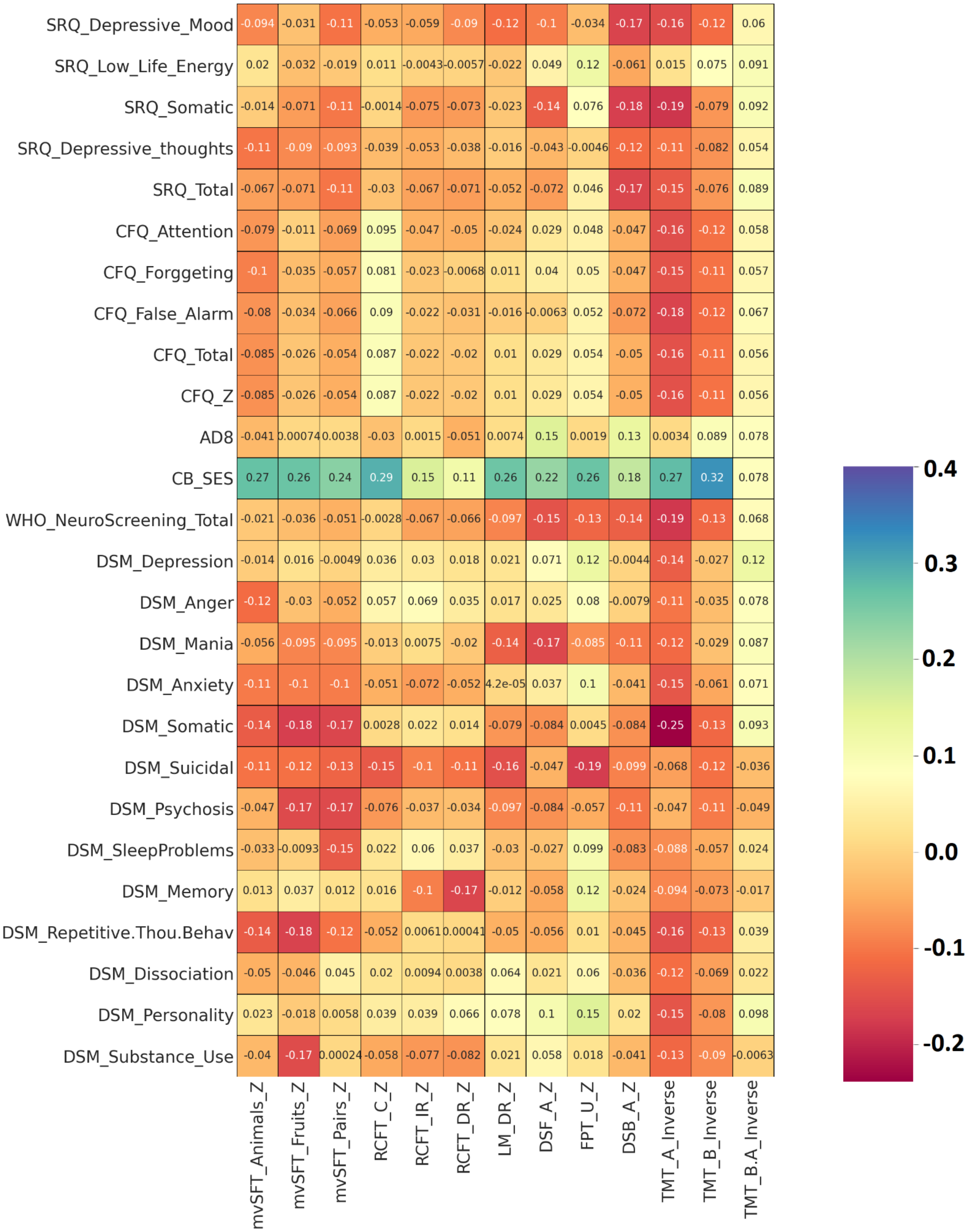


Spearman correlation was performed using the Pandas library in Python3. The correlation product was plotted on a heatmap using the SeaBorn library. The y-axis of the image shows attributes of the questionnaires considered for the correlation analysis, and the x-axis shows the results of neuropsychological tests. No strong correlations were found, except a few trends regarding socioeconomic status according to the Criteria Brazil of economic classification ("Critério Brasil - ABEP" n.d.).
